# Contributing Factors to Dietary Quality and Food Security in Low-Income Households with Children in the United States: A Scoping Review

**DOI:** 10.1101/2022.09.06.22279548

**Authors:** Denise Mc Keown, Lisa Graves, Bethany McGowan, Heather A. Eicher-Miller

## Abstract

**Background:** Low income and food insecure households are at risk of poor dietary quality and food insecurity. Especially in childhood, consuming a nutritionally adequate diet is an essential driver of health, growth, and development. Prior research has shown many household-level factors can present challenges to support the nutritional needs of the members of low income and food insecure households.

**Objective:** The aim of the scoping review is to identify the contributing factors to dietary quality and food security in U.S. households of school-aged children and synthesize the evidence.

**Methods:** The scoping review was conducted following the Preferred Reporting Items for Systematic Reviews and Meta-Analysis Protocols Extension for Scoping Reviews (PRISMA-ScR) using search terms addressing food insecurity, low-income and dietary behaviors in the database PubMed (NCBI). Screening by 3 independent reviewers at the title, abstract, and full study phases identified forty-three studies included in the review.

**Results:** The studies addressed six themes: parental behaviors, child/adolescent behaviors, food procurement behaviors, food preparation behaviors, and psychosocial factors. Most studies were cross-sectional (n = 40, 93%) and focused on parental behaviors (n = 24, 56%), followed by food procurement behaviors, and food preparation behaviors.

**Conclusion:** The findings can be used to inform the development of future nutritional education interventions aimed at improving the dietary quality and food security in households with children. The themes identified were interrelated and suggest that providing parents with education on the following topics: 1) the importance of modeling positive eating behaviors in the home, 2) approaches to support and encourage positive feeding practices with their children, and 3) practical strategies to overcome barriers to purchasing and preparing foods of high nutrient quality. For example, delivering educational sessions on meal selection and preparation and improving nutritional knowledge hold promise to improve dietary quality among food insecure and low-income households.

## 1 Introduction

### 1.1 Food insecurity and poor dietary quality in low-income households

About 10.5% of households, or 13.8 million households, within the U.S. experienced food insecurity at some time throughout 2020^1^. Food insecurity refers to a situation where households experience uncertainty regarding having sufficient food or the inability to obtain sufficient food to meet the needs of all family members due to insufficient money or resources^1^. The United States Department of Agriculture (USDA) classifies food insecurity into two categories, low food security and very low food security. Low food security refers to ‘reports of reduced quality, variety, or desirability of diet and little or no indication of reduced food intake’. Very low food security refers to ‘reports of multiple indications of disrupted eating patterns and reduced food intake’^2^. Several factors including income, employment, and ethnicity can influence food insecurity. Insufficient or no money to purchase adequate food is a risk factor for food insecurity^3,4,5^. In 2020, 35.3% of low-income households were food insecure^2^. Both low income and food insecure households are at risk of reduced dietary quality when compared with households with higher incomes^4,6^.

Dietary quality refers to the quality of an individual’s overall food intake^7^. The U.S. as a population do not meet dietary recommendations for fruit, vegetables, and whole grains and exceed dietary recommendations for added sugars, sodium, refined grains, and saturated fat^8,9^. In the U.S., the Healthy Eating Index (HEI) can be used to rate dietary quality as compared with the recommendations of the Dietary Guidelines for Americans (DGA)^10^. With an ideal HEI score of 100 indicating full adherence to the DGA, the average HEI score for Americans is 59. Children between the ages of 6 and 17 years have an HEI score of 53, and adults between the ages of 18 and 64 years have an HEI score of 58^11^. In the HEI-2010 report, food insecure households scored lower than food secure households, with HEI scores for the food at home purchase basket of 44 and 49 respectively^12^. HEI scores may be lower in food insecure households due to irregular dietary patterns, such as skipping meals and eating less or more of certain components than intended or required for health^13^. Households experiencing food insecurity and households with low-income report increased intake of sugar sweetened beverages, red/processed meat, salty foods and full fat dairy products and lower intakes of fruit, vegetables and fiber which may contribute negatively to dietary quality^10^. Poor dietary quality among low-income and food insecure households is associated with increased prevalence of health conditions, including some cancers, type 2 diabetes mellitus, obesity and cardiovascular disease^14,15,16^. Furthermore, food insecurity has been associated with difficulty managing existing health conditions^16^.

Along with reduced dietary quality, food insecurity is also linked to increased risk of nutrition insecurity^17,18^. According to the U.S. Department of Agriculture (USDA), nutritional security refers to “a situation that exists when all people, at all times, have physical, social, and economic access to sufficient, safe, and nutritious food that meets their dietary needs and food preferences for an active and healthy life’’^17^. Nutritional security addresses diet quality as well as food access, emphasizing the relationship between food insecurity and an increased risk of diet-related illnesses^19^. Poor nutrition is noted as a leading cause of illness in the U.S., accounting for more than 600,000 deaths per year, or more than 40,000 each month^20^. Greater access to healthy meals and improved assistance would help reduce nutrition insecurity in the U.S., with the goal of achieving optimal physical and mental health among the U.S. population^18^.

Specifically for children, consuming a nutritionally adequate diet is an important driver in health, growth, development, and wellbeing^21^. Childhood is a special time of physical, mental, and emotional development where specific amounts and types of nutrients are necessary at critical times for achieving full genetic potential. Dietary behaviors are established in childhood and continue into adulthood^22^. Food insecurity experienced during childhood can be associated with the consumption of a nutritionally inadequate diet and poor health outcomes^23^. According to the HEI-2015, dietary quality was lower in food insecure children compared to food secure children^24^. Children who are food insecure are more likely to have poor overall health, twice as likely to have asthma, and are nearly three times more likely to have iron deficiency anaemia than food-secure children^25^. Children in households with low incomes also have poor dietary quality and increased prevalence of poor health conditions, including obesity, mental health problems, and asthma compared with children from households with higher incomes^26^. The highest prevalence of childhood obesity was among children from families with low incomes^27^. Studies investigating the nutritional quality of children’s diet in relation to family income concluded that children from low-income households had lower intakes of dietary fiber, calcium, fruit, and whole grains along with higher levels of sodium, refined sugars, and saturated fat^28,29,30^.

### 1.2 Limited resources constrain dietary quality and impact relationships among low income and food insecure households

Both low income and food insecure households can face challenges in obtaining food to support the nutritional needs of family members. Availability of food resources has been hypothesized as a major determining factor for purchasing a high-quality diet^31^. Along with economical restraints, dietary choices in family households can be influenced by multiple challenges including availability, accessibility, food preferences, eating behaviors, parental nutritional behaviors and knowledge, parental modeling and psychosocial precursors^10,22^.

Several studies have examined the association between income and dietary quality and food security highlighting income as the most significant challenge for food insecurity and dietary quality in households with low incomes^10,31^. The cost of healthy food is an economic driver of food insecurity and poor dietary quality. Regardless of individuals having the necessary knowledge and skills, a limited budget may result in individuals opting for food of best economical value instead of food with the most nutritional value. Studies examining food purchases and nutritional quality concluded that households with lower incomes purchased foods of lower nutritional quality when compared to households with higher incomes^10,12,32^.

Parental influence, including nutritional beliefs, attitudes, and knowledge play a role in the dietary quality of their children’s diets as well as their children’s health, nutritional knowledge, and eating behaviors^33,34^. Furthermore, it is important to understand the distribution of food between members of the family in order to assess the dietary quality of the entire household. In food insecure households with children, parents may protect their children from negative changes to dietary quality and quantity by using strategies such as limiting or altering their own dietary intake^35^. In 2020, 14.8% of households with children experienced food insecurity. However, in half of these households, only the parents experienced food insecurity^1^. Therefore, food insecurity and poor dietary quality may be more evident in adults than in children. Furthermore, food security may also differ by age of the child. Adolescents are twice as likely to experience food insecurity when compared with younger members of the family. This may be because adolescents are more independent from the household and adolescents may be able to access food outside of the home more easily^37^.

### 1.3 Rationale to investigate the factors influencing dietary quality and food security among low-income households

Food expense, family relationships, and other recognized influences described above are known to impact dietary quality and food security in low-income households. Yet, interest in additional factors influencing dietary intake and food security has grown since the greater recognition of food security in the 2010 DGA and more recent recognition of nutrition security^38,19^. Furthermore, knowledge of both parents’ and children’s behaviors, food procurement and preparation behaviors, and psychosocial aspects spanning a wide range of studies and literature are important in gaining a broader understanding and synthesis of the relevant scientific evidence. A scoping review can be used to comprehensively identify and integrate the factors influencing dietary quality and food security in low-income households as well as to identify any existing gaps in knowledge. Understanding families’ priorities and challenges in the purchasing and consumption of nutritious foods is important for informing and implementing future nutritional interventions targeted at improving dietary quality in low income and food insecure households. A scoping review can be used to integrate prior knowledge to maximize the impact of future nutritional education interventions in households^39^.

This scoping review aims to determine the contributing factors to dietary quality and food security in low-income U.S. households with children. The purpose of this review is to synthesize the evidence on this topic and identify the factors affecting the dietary quality and food security of low-income households with children.

## 2 Methods

### 2.1 Literature search strategy

The PRISMA (Preferred Reporting Items for Systematic Reviews and Meta-Analysis Protocols) Extension for Scoping Reviews (PRISMA-ScR) was utilized to conduct the review^40^. Search strategies were developed collectively through discussion between four researchers. The following search term was used: (food insecurity OR food security OR low-income OR poverty) AND United States AND (dietary quality OR dietary behavior OR dietary selection OR dietary attitudes OR family meal planning OR food purchasing). The online database, PubMed (NCBI) was used to search for studies published within the last 10 years between 2012 and 2022.

The inclusion criteria were as follows: all studies written in English and published within the last 10 years (2012-2022) which focused on food security and/or dietary quality of low-income families in the U.S., and the factors/behaviors/attitudes/barriers affecting food security and/or dietary quality in these households.

Excluded studies were those that did not match the conceptual framework of the study, not written in English, carried out outside of the U.S., not focused on low-income or food insecure households, those focused on households without children and those not focused on school aged children.

### 2.2 Study selection

The PubMed (NCBI) search identified 2,127 studies. The search results were exported to EndNote and Rayyan software, which was used to remove duplicates. Three duplicates were removed. Studies were reviewed at 3 stages: title, abstract and full text. Each stage was reviewed by 2 independent reviewers. Firstly, titles and abstracts were screened based on eligibility criteria. Titles and abstracts were screened by two reviewers independently. Studies were either marked as ‘included’, ‘excluded’, or ‘maybe’ by the reviewers. An independent reviewer was a tiebreaker who reviewed the titles and abstracts of studies in instances when there was conflict on study inclusion or exclusion. Any studies that were judged as ‘maybe’ were treated as ‘included’. Next, the remaining chosen studies were screened by reading the full text using the same process as the screening for previous stages, using the inclusion criteria. One hundred and seven studies were selected for full-text evaluation. Forty-three studies were included in the scoping review (Figure 1).

**Figure 1.**
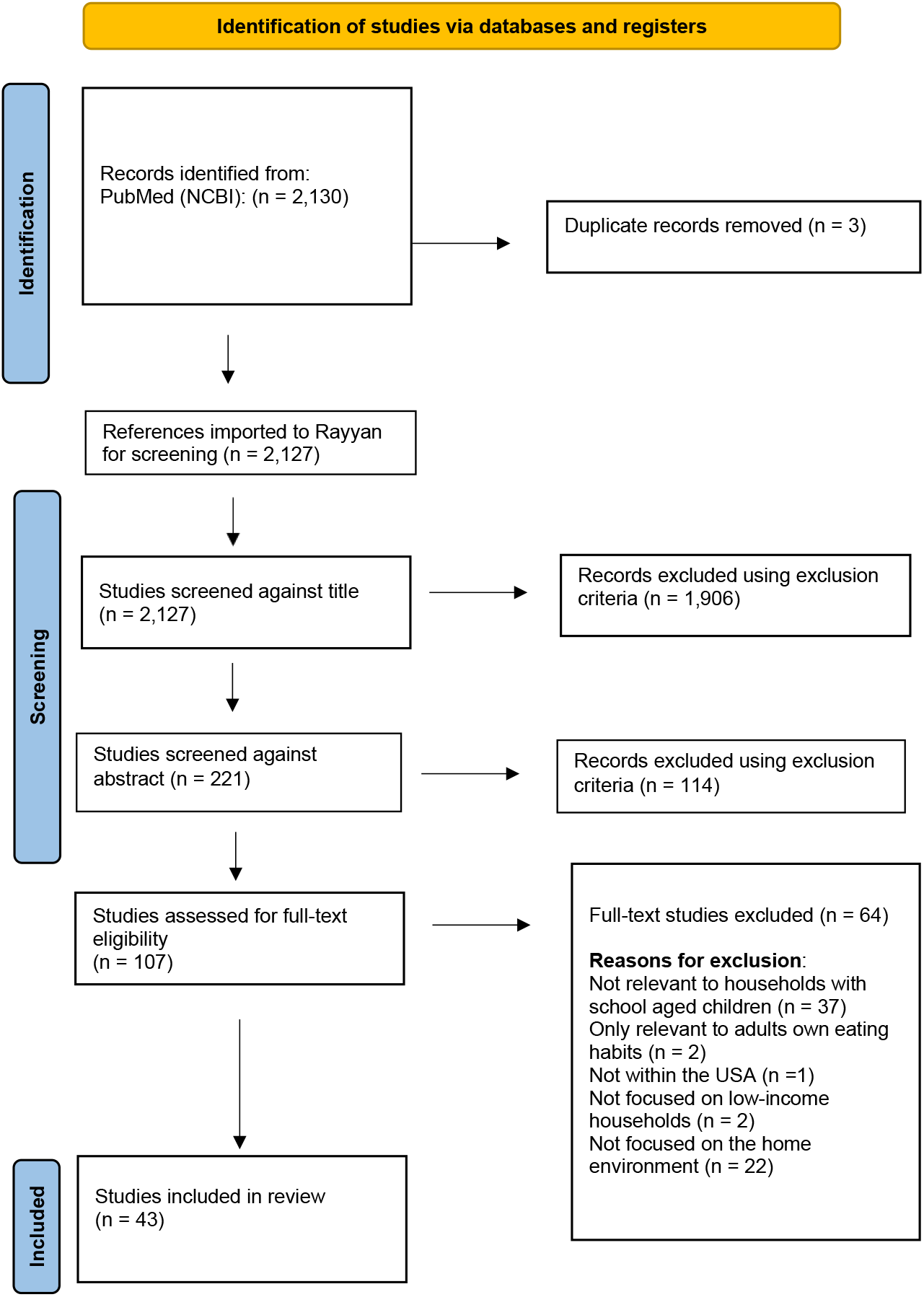
PRISMA flow diagram

### 2.3 Data synthesis

The studies’ title, author, aims, study design, studied population, and main findings are presented in **Table 1**. Most of the 43 studies were cross-sectional (n = 40, 93%), followed by longitudinal (n = 1, 2%) and systematic reviews (n = 2, 5%). Contributing factors to dietary quality and food insecurity in low-income households were organized into five themes: 1) parental behaviors, 2) adolescent/child behaviors, 3) food procurement behaviors, 4) food preparation behaviors, and 5) psychosocial factors. Several studies addressed more than one of the themes. Twenty-four studies (56%) included parental behaviors, 7 studies (16%) included adolescent/child behaviors, 13 studies (30%) included food procurement behaviors, 12 studies (28%) included food preparation behaviors, and 11 studies (26%) included psychosocial factors. **Table 2** identifies the themes discussed in each study.

**Table 1.**
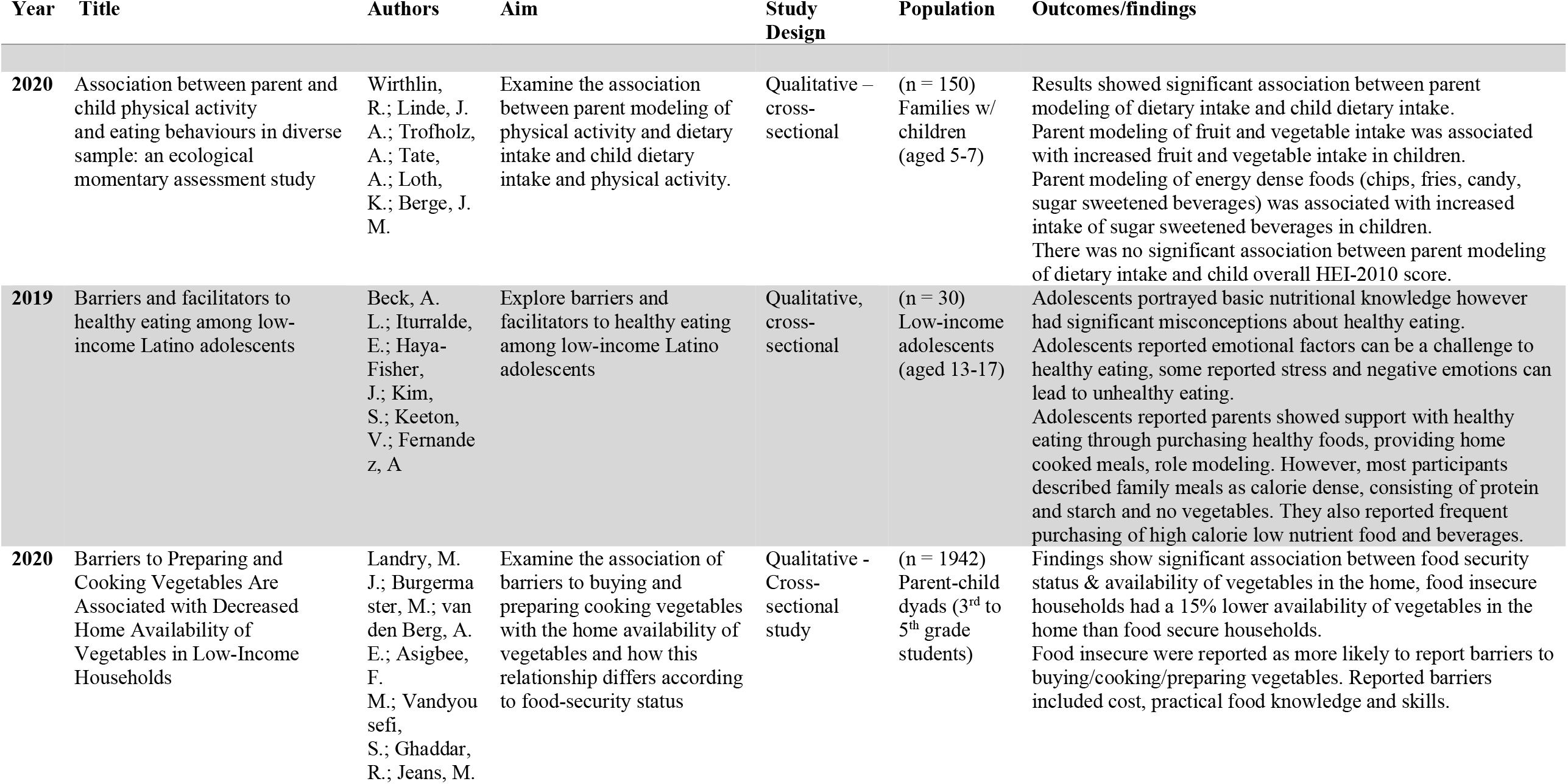
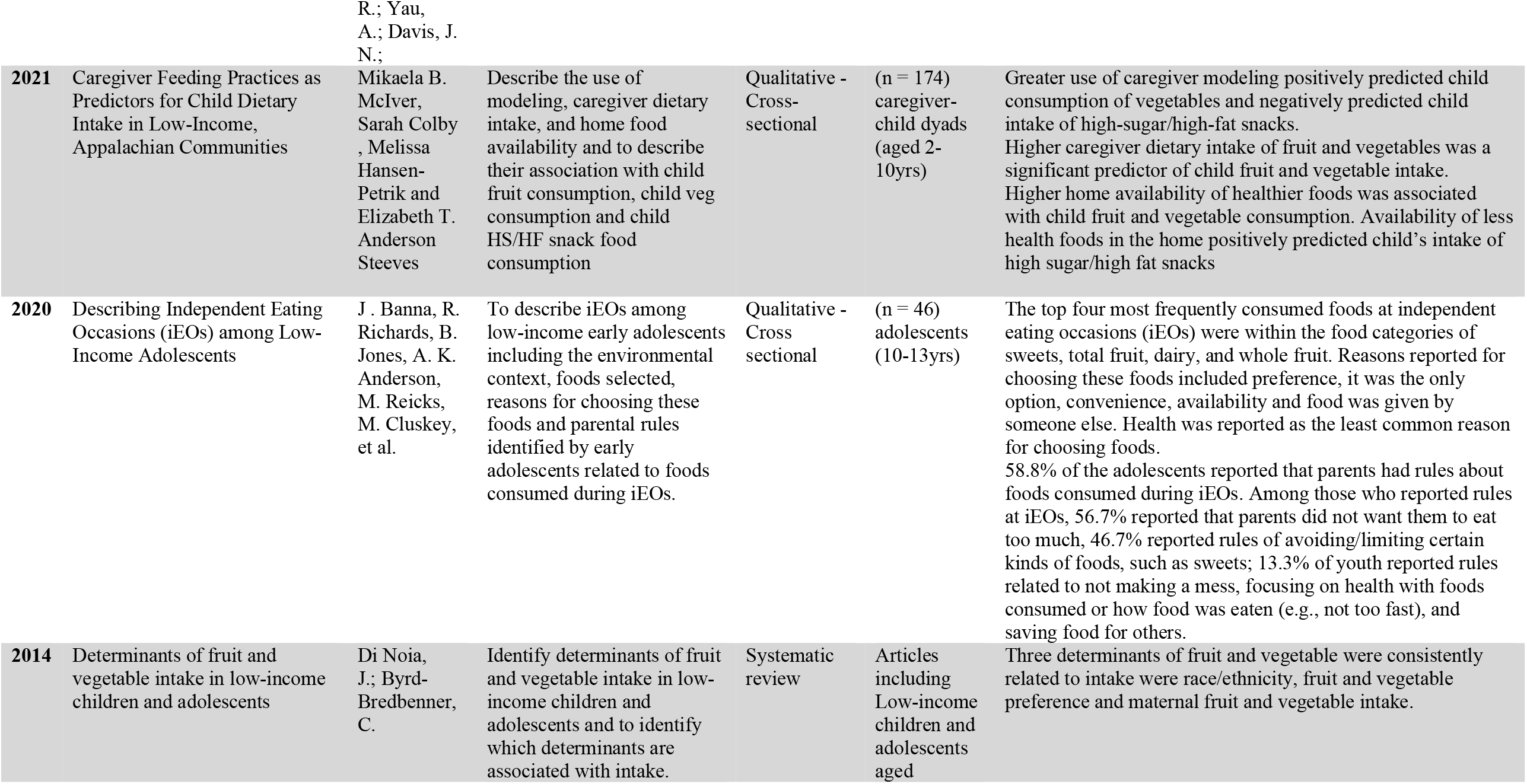
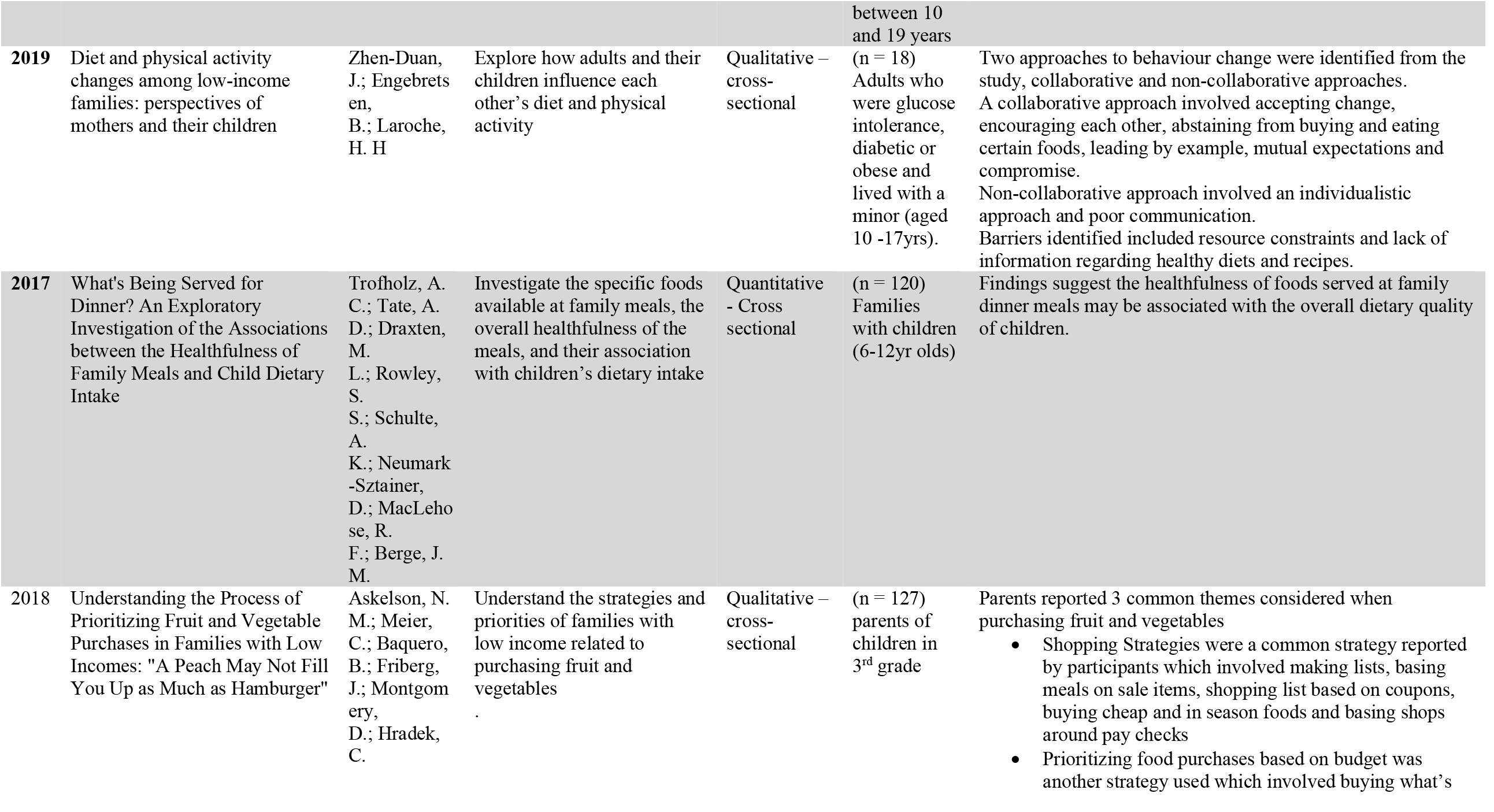
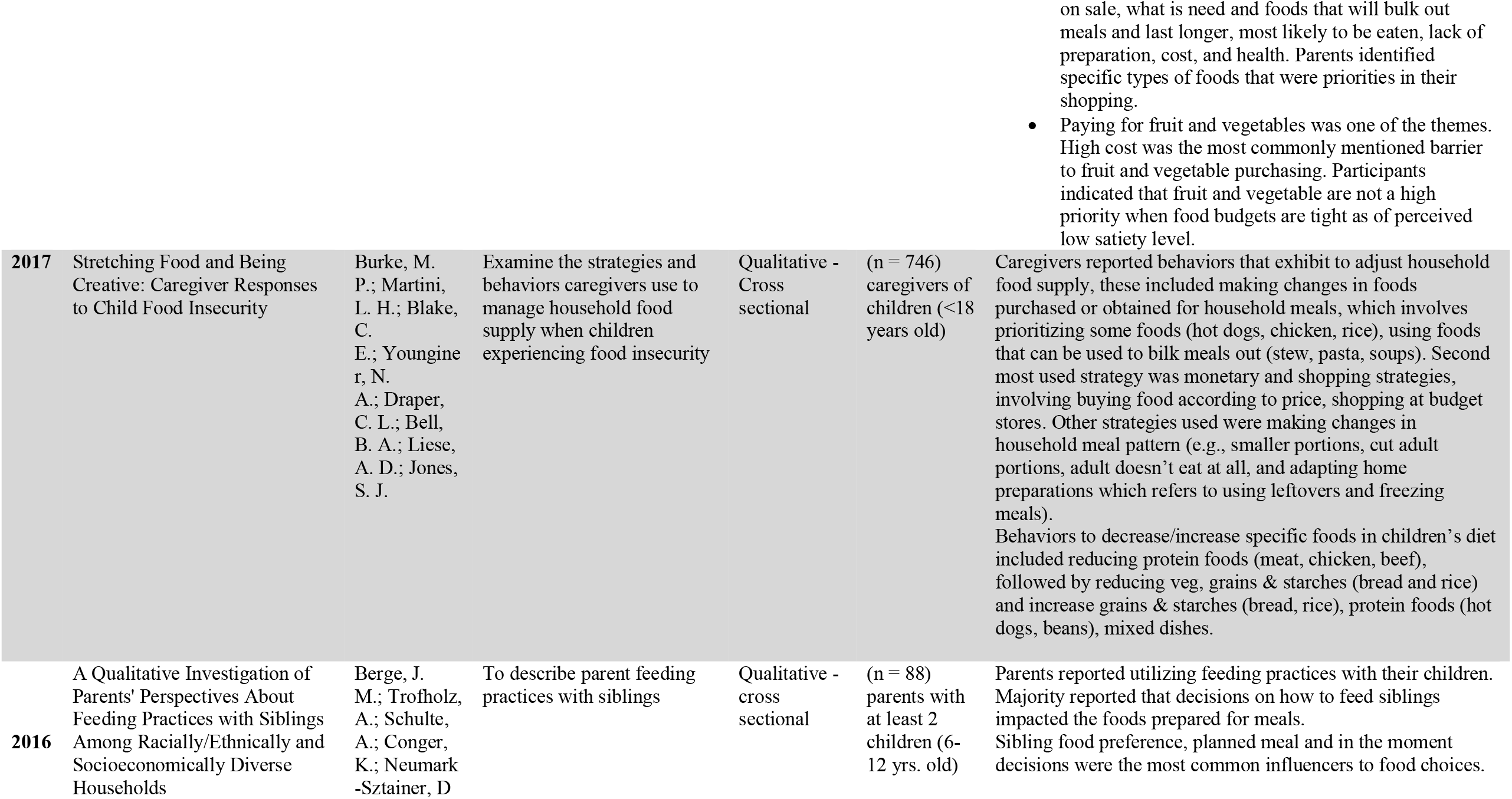
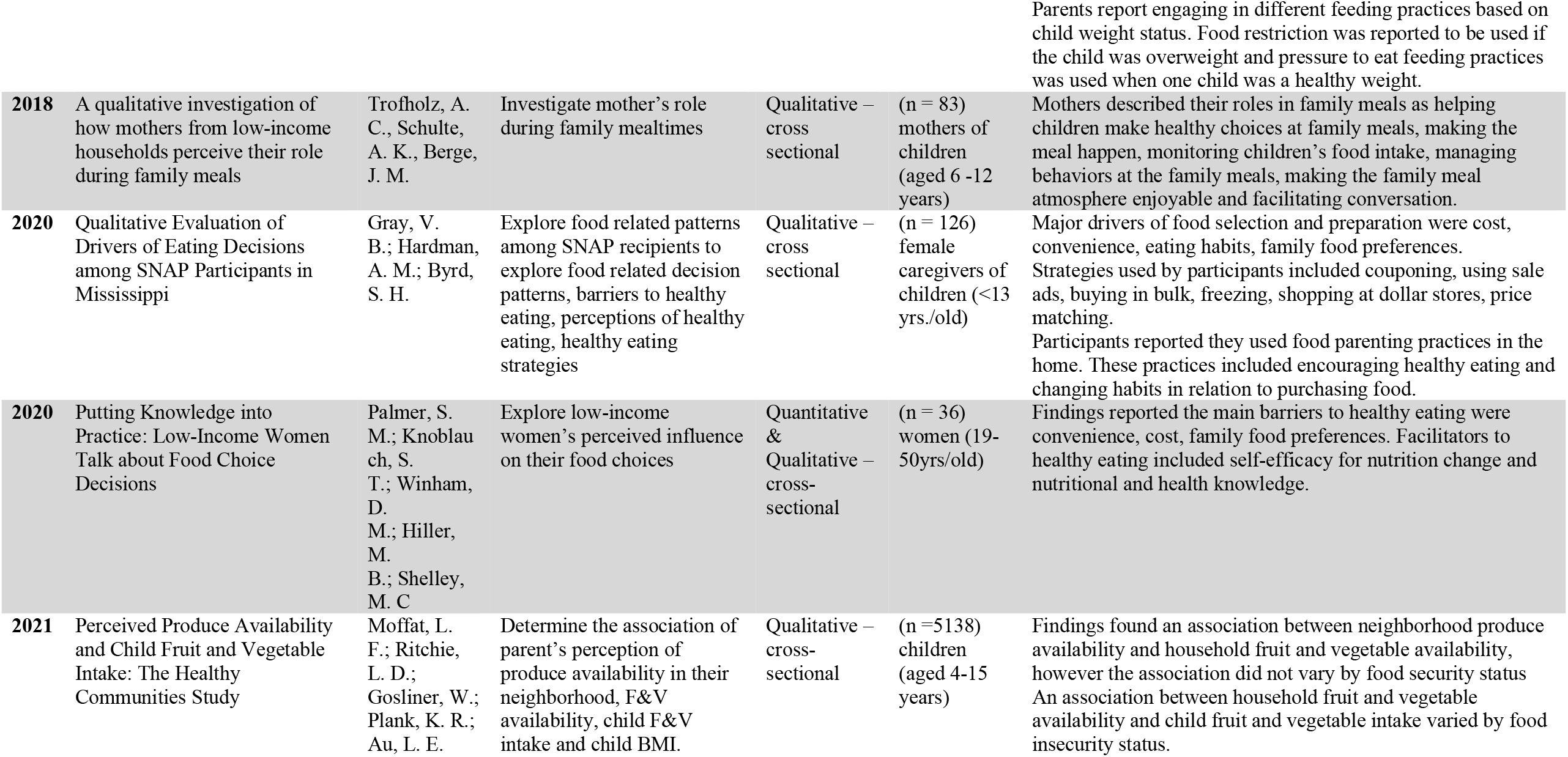
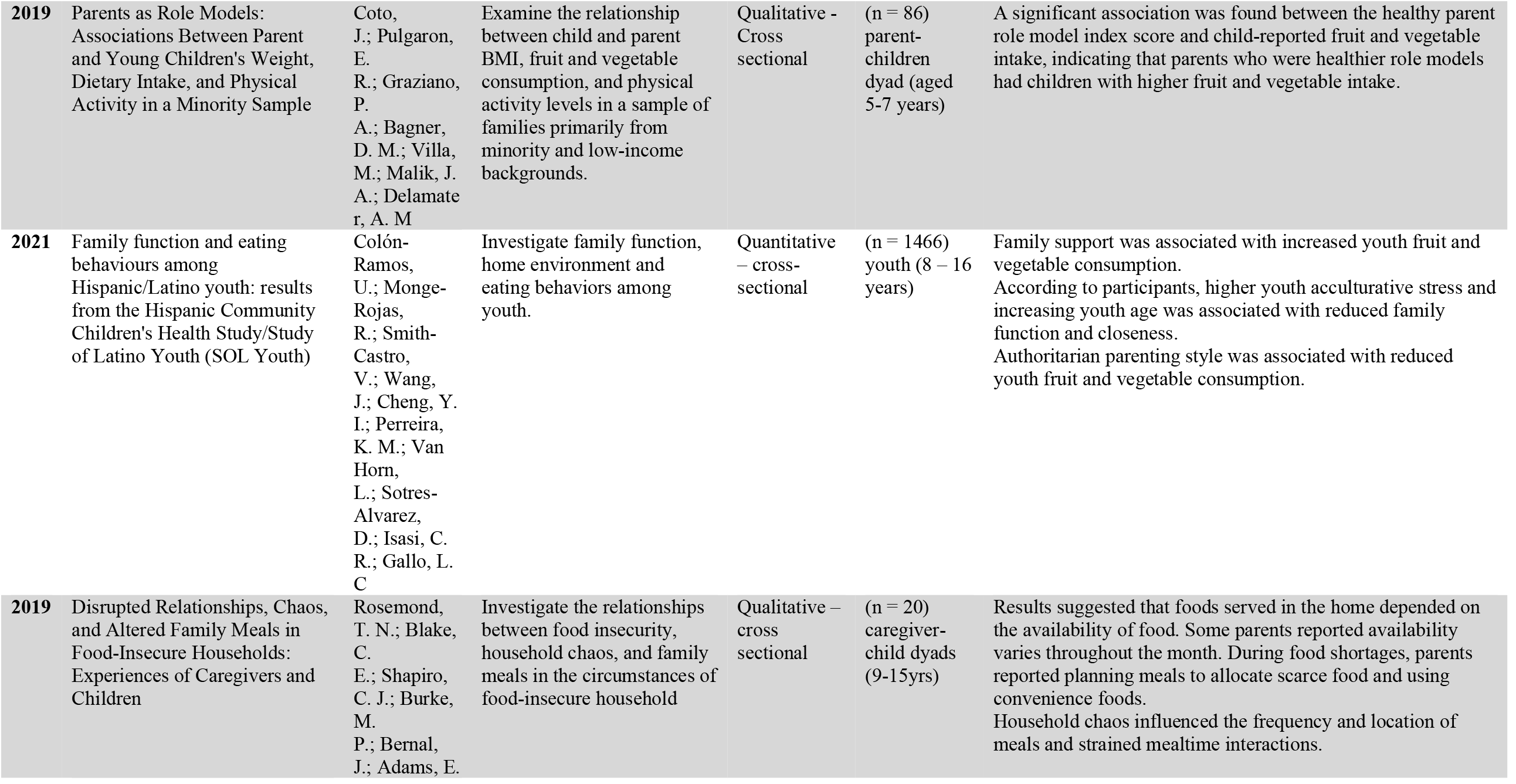
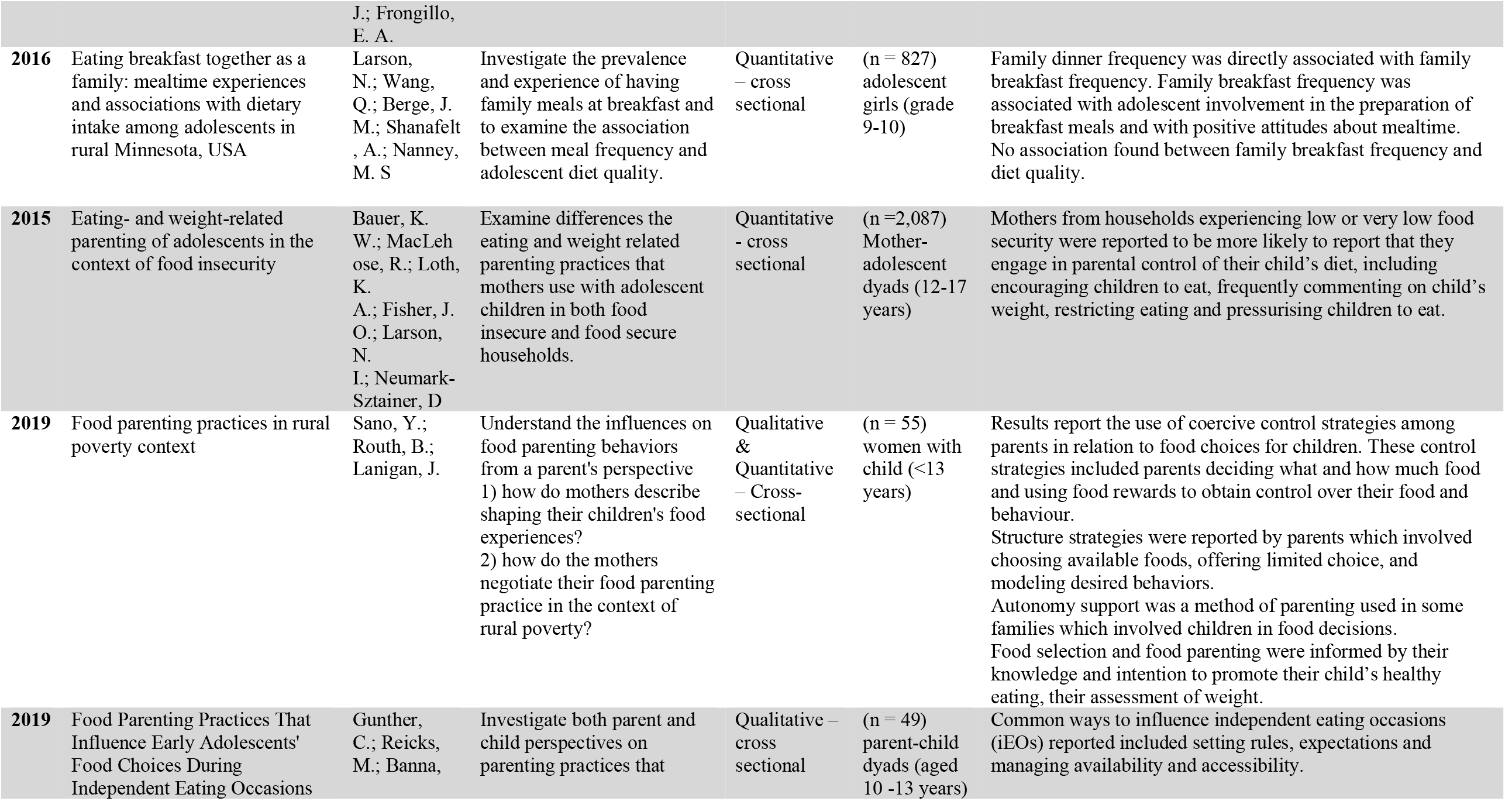
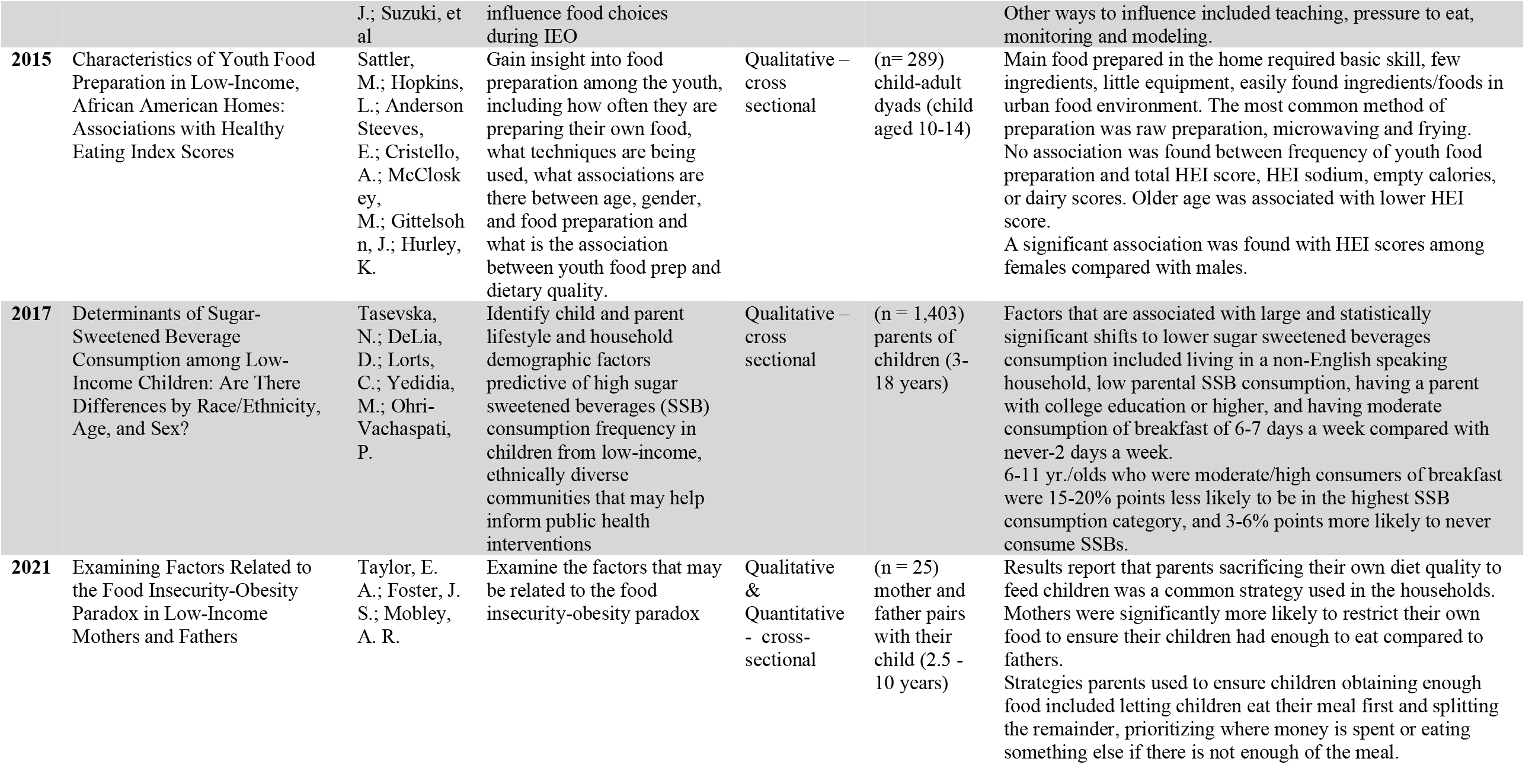
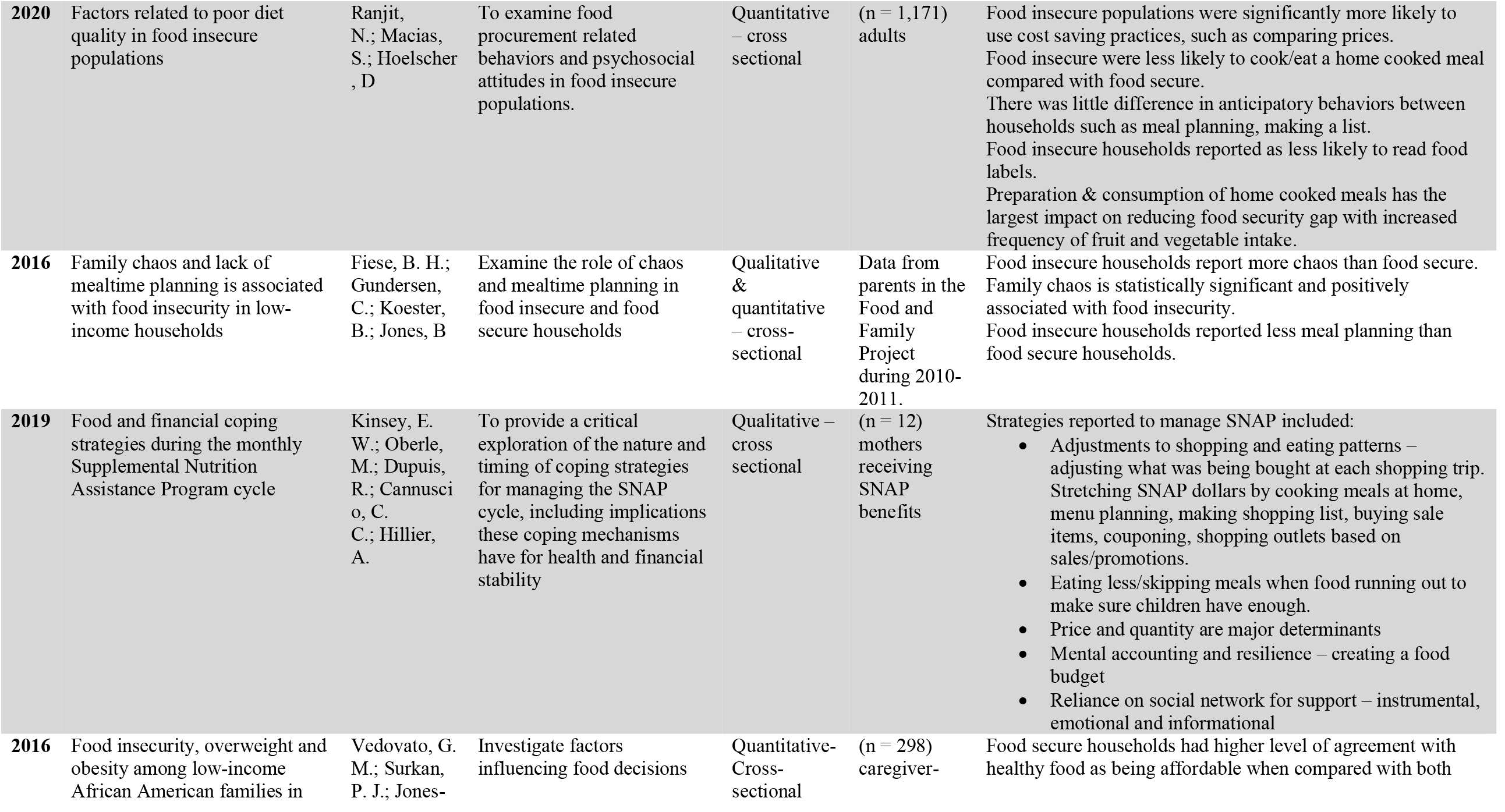
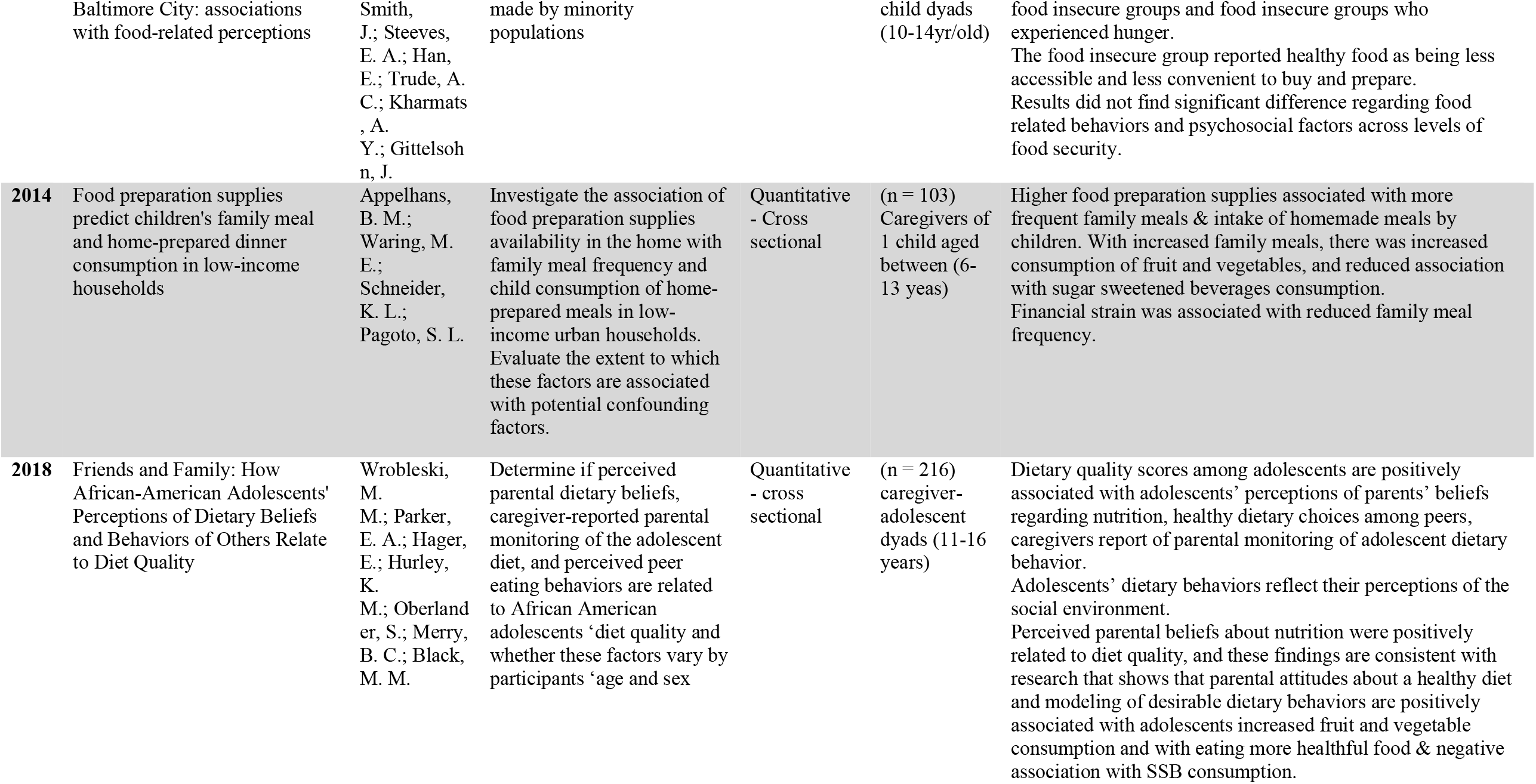
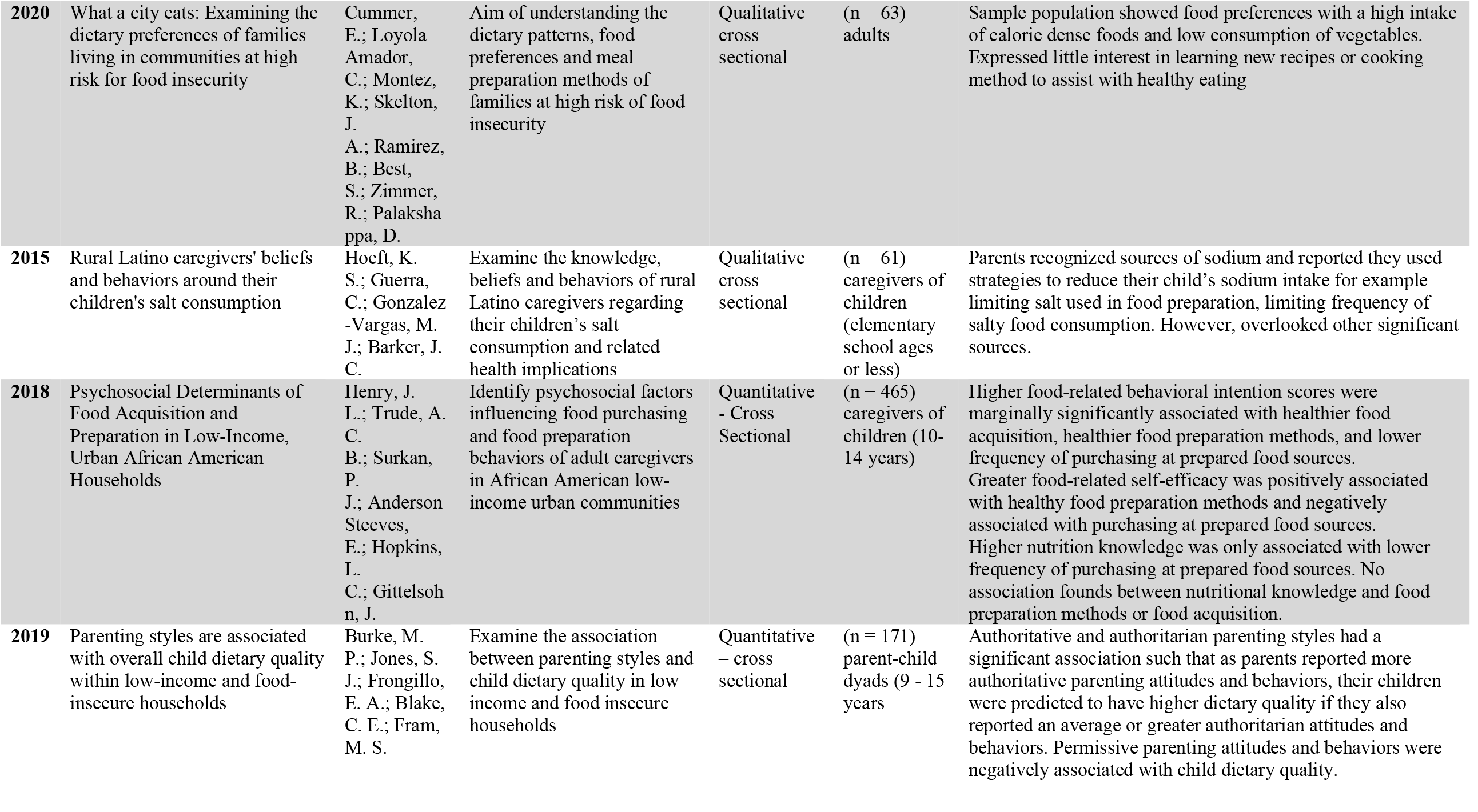
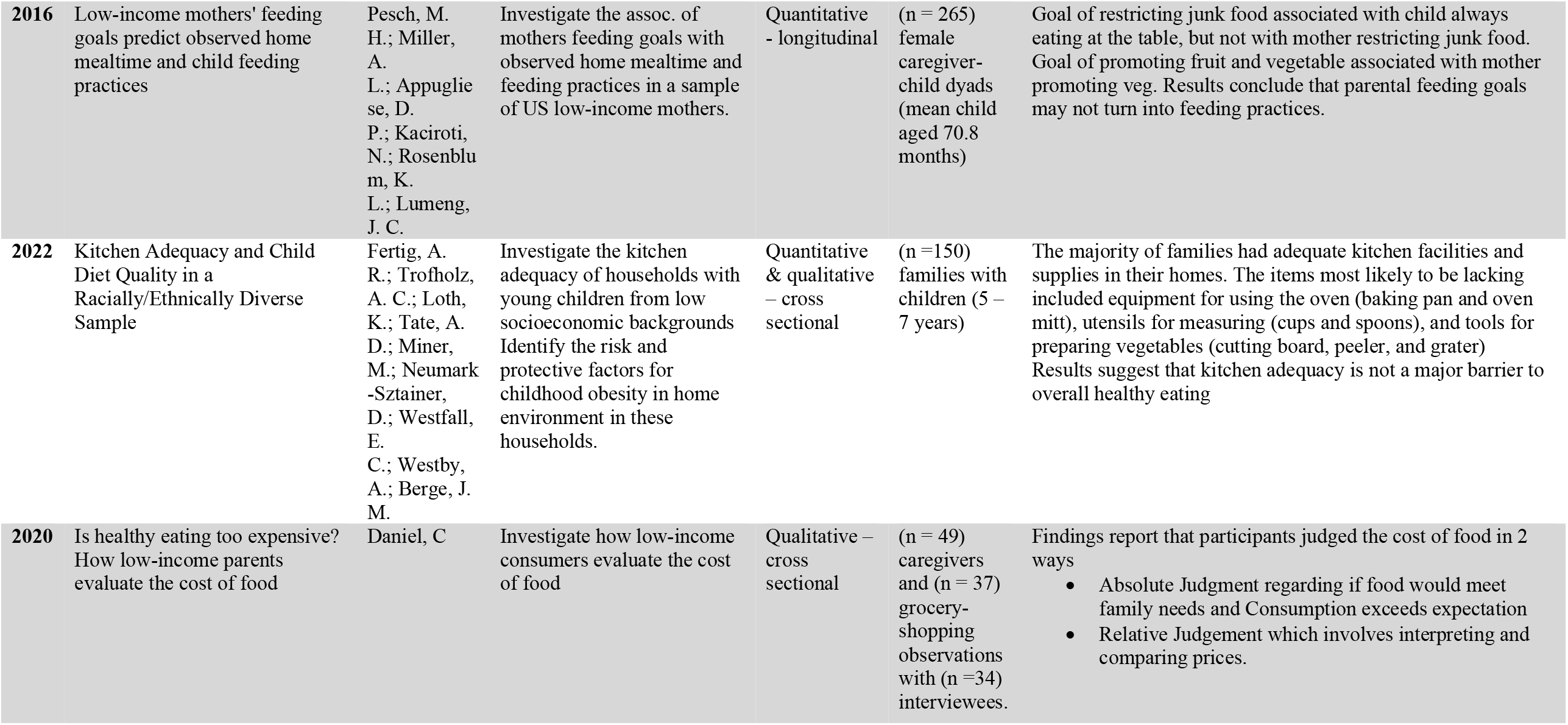
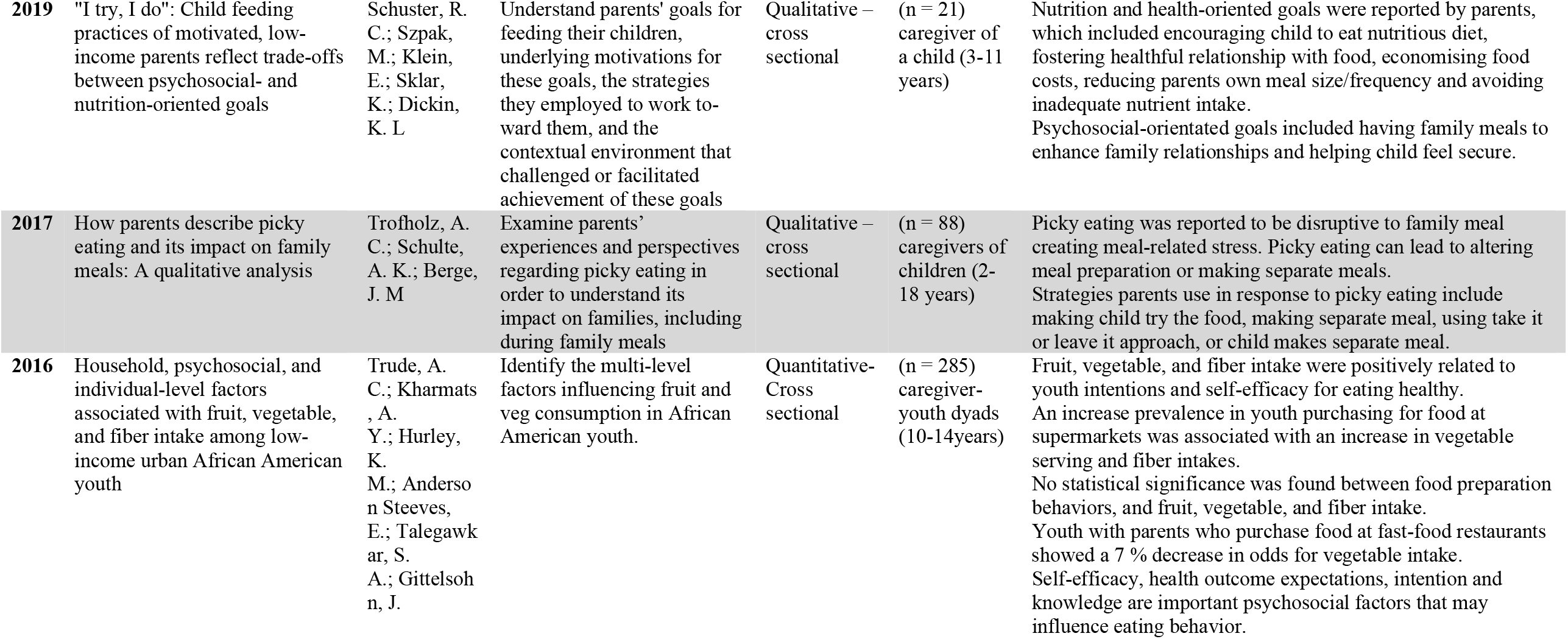
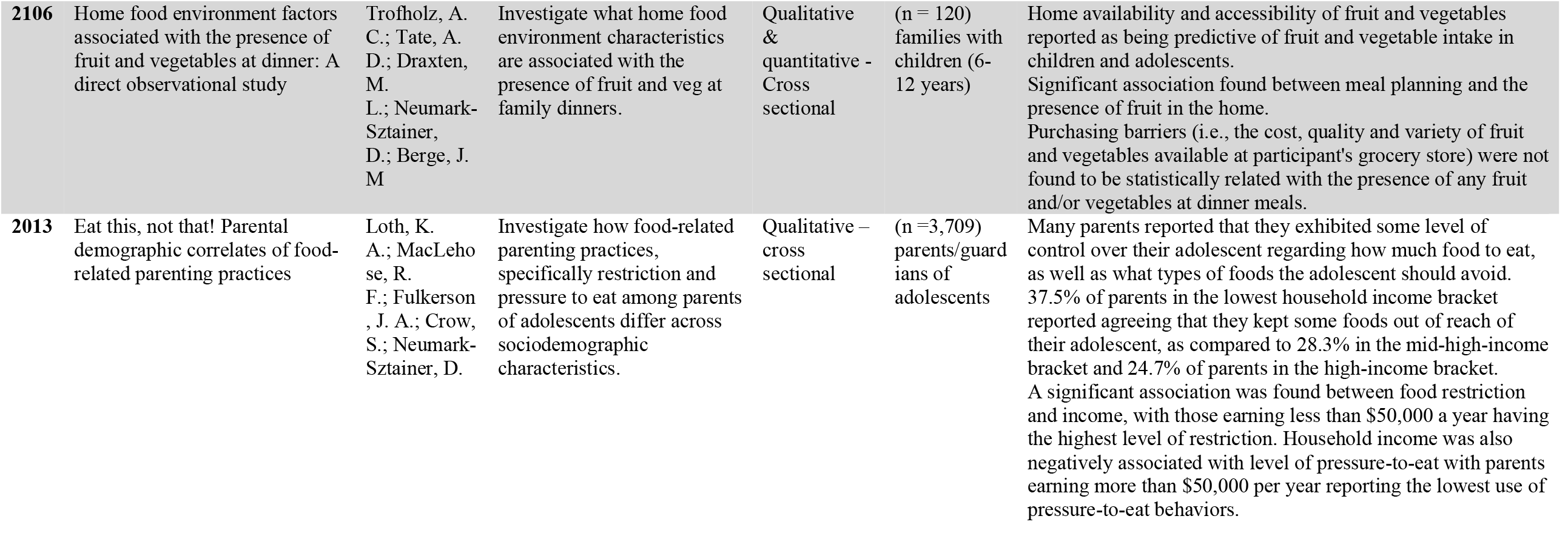
Characteristics of sources of evidence and main findings

**Table 2.**
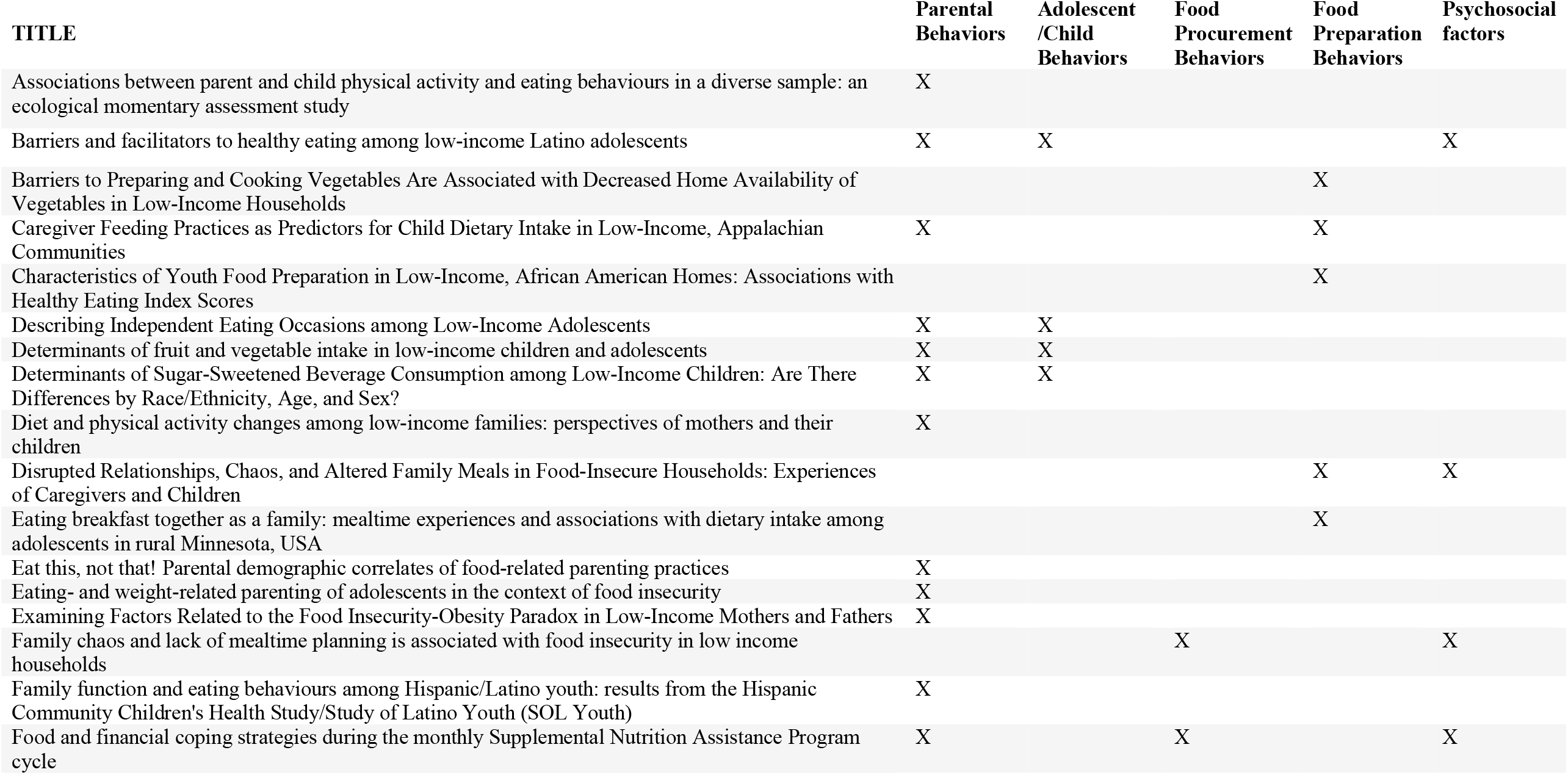
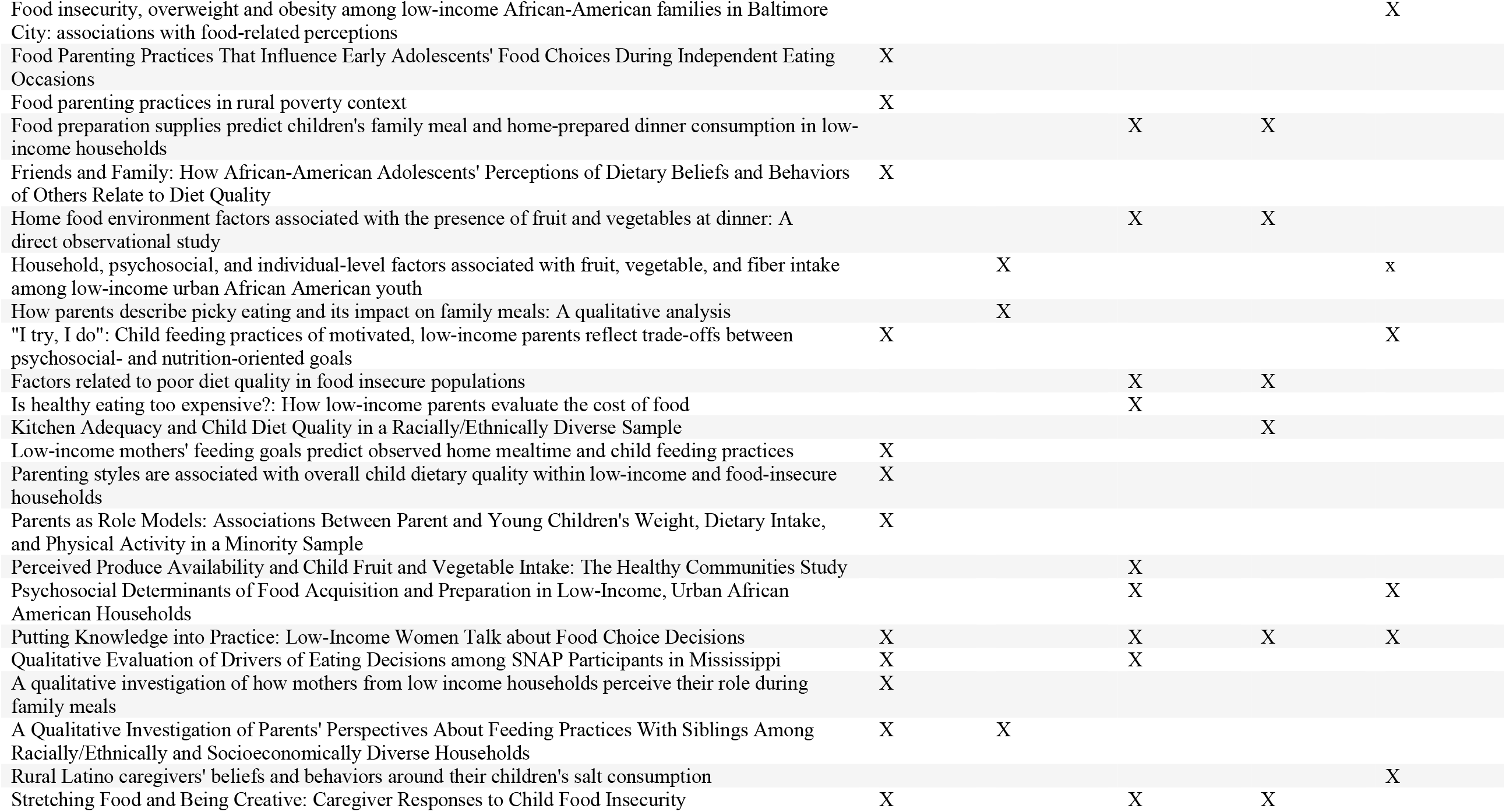
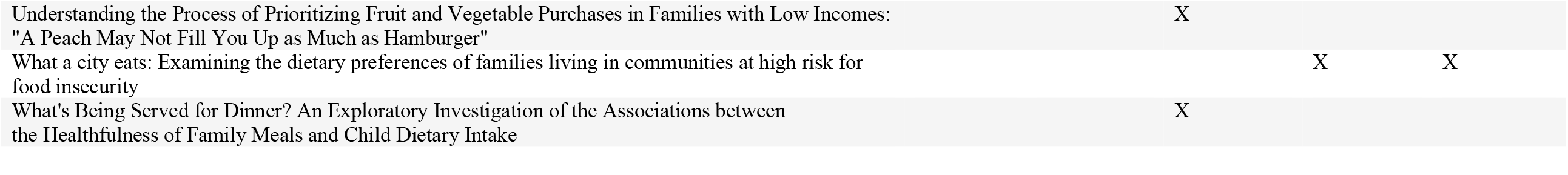
Themes identified in each source of evidence

## 3 Results

### 3.1 Parental behaviors

Parental behaviors investigated included parental modeling, feeding practices, parental attitudes and support, and parenting styles. Parental modeling, in 11 of the studies^41,42,43,44,45,46,47,48,49,50,51^, refers to a parent’s effort to demonstrate healthy food choices and healthy eating behaviors with the objective that the child will exhibit similar behaviors. Parental modeling was associated with shaping child’s eating behaviors^41^. Four studies showed an association between parent modeling and increased fruit and vegetable intake among children while modeling of energy-dense food was associated with increased sugar sweetened beverage intake^42,43,44,45,46^. One study reported healthy eating modeling was associated with reduced child consumption of high-sugar/high-fat snacks^45^. In 5 studies, parental modeling acted as a support for encouraging healthy eating behaviors among children^47,48,49,50,51^.

Parental feeding practices linked to child behavior were investigated in several studies^50,51,52,53,54,55^. Parents reported engaging in different feeding practices to influence children, such as pressure to eat, food restriction, and controlling food intake, based on child characteristics, including weight, age, and developmental stage^50^. According to three studies, parents engage in behaviors to control the feeding environment in the home, such as food restriction feeding practices when the child is overweight and pressure-to-eat feeding practices when the child is a healthy weight^50,51,52^. According to two studies, income and food security are linked to food restriction and pressure-to-eat behaviors, with low-income and with low or very low food secure households being more likely to engage in these food restriction and pressure-to-eat practices^53,54^.

Parental eating attitudes and support from parents play a role in eating habits among children. Parents reported encouraging healthy eating behaviors through practices such as limiting the availability of sugar-sweetened foods in the home. On the other hand, the same study indicated that parents buy foods that they know their children will eat to avoid buying foods that will go to waste, which can contribute to the purchasing of foods with reduced dietary quality^55^. Parents reported monitoring their children’s food intake, establishing their own healthy relationship with food, and encouraging their children to select nutritious options to assist them in making healthy food choices^55,56^. Parents also had rules in place for adolescent dietary intake, such as limiting particular foods in the home, and focusing on healthy foods^57^. One study examining breakfast frequency found that family meal frequency was associated with positive attitudes around meals and food^58^.

Parents can engage in strategies to control the home feeding environment with the aim to control their child’s intake of foods, like adapting the type and amount of food served at meals^59,60^. In food insecure situations, parents reported adapting their own dietary intake with the aim of their children receiving sufficient amounts of food, by reducing the parent’s own portion sizes and skipping meals^56,59,60, 61,62^. Parents from low-income households also expressed goals to restrict junk food in the home, encourage the child to eat a nutritious diet, and to promote fruit and vegetable intake^56,63^. However, one study also noted that not all parental feeding goals turn into practices in the home^63^.

Studies^34,61^ on parenting styles and impact on dietary quality of children were identified. One reported that parents who adapted authoritarian parenting attitudes and behaviors were positively associated with dietary quality of the child. Authoritarian parenting attitudes and behaviors typically involve parents having high demands, low responsiveness, low emotional warmth, and unwillingness to negotiate. A permissive parenting style, having few rules and giving children freedom in choice, was negatively associated with child dietary quality^61^. Alternatively, the second study indicated an association between authoritative parenting style and reduced fruit and vegetable consumption among adolescents^34^. Parenting styles should be considered as a potential determinant that is associated with overall nutritional quality.

### 3.2 Adolescent/Child behavior

Adolescent/Child behaviors in 9 studies^41,43,46,52,57,59,65,66,67^, included investigations of personal food preferences, healthy eating attitudes and behaviors, and picky eating. Adolescents reported choosing food based on preference, convenience, the foods available in the home or the food offered to them by someone else. Health was considered the least common reason for choice^57^. Adolescent preference was noted as a determinant in adolescent dietary intake in two other studies^41,57^. According to one study, adolescent fruit, vegetable, and fiber intake was positively associated with healthy eating intentions and self-efficacy^65^. Eating attitudes and behaviors were associated with dietary quality^46^.

Adolescents demonstrated basic nutritional knowledge and were able to recognize healthy and unhealthy foods. However, the adolescents held some misconceptions around healthy foods^47^. One study reported that youth self-efficacy, nutritional knowledge and healthy attitudes were important psychosocial factors that influence youth eating behaviors^65^. Another concluded that psychosocial factors, including knowledge and self-efficacy, did not differ between food secure and food insecure households^67^.

Adolescents also reported being motivated to adopt healthy dietary practices in their diets however some reported that it was difficult to sustain these healthy eating changes^47^. Picky eating among children was disruptive on family mealtime and the home environment, causing meal-related stress and increased time parents spend in meal preparation as picky eating can require parents to adjust or make additional meals^52,66^.

### 3.3 Food procurement behaviors

Food procurement behaviors in 9 studies^13,55,59,60,61,68,69,70,93^ comprised of topics including purchasing behaviors and strategies and barriers to purchasing foods. Priorities that individuals considered when purchasing foods included cost, family preferences, food preparation time, family needs, and shelf life. Studies reported behaviors engaged in when purchasing foods included making lists, buying sale items, comparing prices, shopping in discount stores, and using coupons^13,55,59,60,61,68,69^. Price was a major determinant in purchasing behaviors^61^. These barriers were more often reported in food insecure households compared to food secure households^70^. One study compared food procurement behaviors in food insecure and food secure households and reported food insecure households being significantly more likely to use cost saving practices, such as price comparisons. However, there was no difference between the groups in the use of anticipatory behaviors, including meal planning and making a shopping list^13^. Two studies reported that the high cost of fruit and vegetables was the main barrier in purchasing^68,70^. In contrast to these findings, one study found no relationship between purchasing barriers and the presence of fruit and vegetables at dinner meals^93^.

### 3.4 Food preparation behaviors

Food preparation behaviors like incorporating meal planning, kitchen/cooking supplies, cooking skills, and food availability were addressed in 12 studies^13,5,59,60,70,71,72,73,74,93,94^. Availability of healthy foods was associated with dietary quality and food security in five of the studies^45,70,71,93,94^. One study reported an association between food security status and the availability of vegetables in the home^70^, while two studies reported an association between availability of fruit and vegetables in the home and increased child consumption of fruit and vegetables^93,94^. Similarly, two studies also suggested that the healthfulness of foods available may be associated with child’s dietary quality, with increased fruit and vegetable intake^45,71^. Consistently, results indicate a relationship between the availability of healthy foods in the home and improved dietary quality and food security.

The accessibility of food preparation supplies was investigated in two studies^72,75^. One study highlighted that availability of food preparation supplies in the home is associated positively with frequency of family meals. Fruit and vegetable consumption increased with frequency of family meals, while sugar sweetened beverage consumption decreased^72^. Contradictorily, a longitudinal observational study’s results suggested that kitchen adequacy is not a barrier to healthy eating^75^.

Several studies investigated meal preparation and planning behaviors^74,94,13,60,73,60^. One study reported food insecure households were less likely to prepare a home-cooked meal compared to food secure households. Furthermore, findings show that low-income households, regardless of food security status, have a low prevalence of planning behaviors related to buying and preparing food^13^. Lack of sufficient time to prepare foods was identified as a barrier to making home cooked meals^60^. The most common food preparation methods required basic skills, little equipment and easily accessible ingredients^73^. Food insecure compared with food secure households were reported to be more likely to report barriers^70^.

### 3.5 Psychosocial factors

Ten studies discussed the influence of psychosocial factors, self-efficacy, nutritional and health knowledge, emotional factors, and family relationships on dietary quality and/or food security^13,47,56,60,76,77,78,79^. Self-efficacy, which refers to a person’s belief in their ability to engage in healthy eating behaviors, was associated with increased dietary quality and healthy eating behaviors in low-income families and adolescents^47,60,76^.

Nutritional knowledge^13,47,77^ was also shown to influence dietary quality. Caregivers had a basic understanding of sodium sources but had difficulty identifying sodium in sources such as cheese and prepared soups. Caregivers had little knowledge of the effects of increased salt consumption on childhood health^77^. According to one study, food insecure adults were less likely than food secure adults to read food labels^13^. According to the findings, knowledge about healthy eating and foods is an important component to focus on in future interventions.

Finally, household chaos was identified as an influencing factor on dietary quality and food security, owing to its negative impact on family relationships and mealtime-related stress^56,79^. Family chaos was significantly associated with food insecurity in one of those studies^79^. A study looking at parenting goals for feeding referred to psychosocial-orientated goals that parents utilized, these included aiming to have family meals to enhance family relationships and helping their child feel secure in the home, which may assist in overcoming mealtime chaos^56^.

## 4 Discussion

This scoping review identified multiple factors, some of which are interrelated, highlighting the complexity of the breadth of evidence assessing key factors to dietary quality and food security in low-income households with children within the U.S. While cost, availability, and family relationships have been identified as key contributing factors, other factors related to dietary behaviors and attitudes, purchasing behaviors, food preparation behaviors, and environmental influences were also notably identified as important to dietary quality in households with low income and households experiencing food insecurity.

Parental behaviors were discussed in multiple studies. Parental behaviors that had the most influence on the household’s dietary quality and food security were parental modeling, parental support and encouragement, parental feeding practices, and parenting styles. Findings highlight the significant influence parents have on the food their child is consuming, including how much and what types of food. Parental modeling of healthy eating was consistent among studies showing a positive impact on their child’s dietary quality. Additionally, parental support and encouragement for healthy eating can improve children’s attitudes towards healthy eating. Parental feeding practices, such as food restriction and pressure to eat, can result in a child’s negative attitude towards food and meals. Overall, parental behaviors should be highlighted with regards to their role in dietary quality and influence on their child’s dietary quality.

Food procurement and preparation factors were commonly noted in this review. Findings suggest that low income and food insecure households experience more barriers to purchasing and preparing foods, including access to food and cost, when compared with households of higher income and food secure households. Overcoming barriers to purchasing and preparing foods may assist in improving the availability of food within the household. The use of strategies in purchasing foods was also mentioned, for example comparing prices, buying sale items, shopping in budget stores, to assist parents in managing their budgets. Results have associated availability of healthy foods in the home with increased dietary quality, and increased consumption of fruit and vegetables, which is consistent with previous studies and reviews^80,81^. In support of this, another study associated the availability of less healthy foods in the home with increased child intake of high-sugar/high-fat snacks^82^, suggesting that increasing the availability of healthful and decreasing less healthful foods in the household may enhance the dietary quality of children.

Lack of nutritional knowledge, along with lack of food preparation and cooking skills, can act as barriers to healthy eating and reduce the dietary quality of foods/meals consumed in the home. Lack of these skills may contribute to the higher prevalence of food insecure groups eating away from home and reduced prevalence of food insecure groups eating home-cooked meals^13^. A previous study investigating the relationship between self-efficacy, with regards to food preparation and cooking, and food security among adolescents concluded that with increased self-efficacy was increased food security^83^. Furthermore, increased nutritional awareness was linked to decreased energy density of the diet and increased food diversity and adequacy in meeting national recommendations across socioeconomic status^84^. The interactions between psychosocial factors, nutritional knowledge and self-efficacy, and dietary quality and food preparation behaviors show the combined influence of personal factors on dietary quality in the home.

## 5 Implications

Federal U.S. nutrition assistance programs, such as the Supplemental Nutrition Assistance Program (SNAP), provide participants with financial benefits to assist with purchasing of food and can play an important role in supporting households to improve dietary quality and food security. In 2021, approximately 42.5 million people, 13% of the population, participated in SNAP^85^. SNAP can assist with increasing accessibility, availability, and intake of heathy foods to improve nutrition security. SNAP has been shown to reduce food insecurity in households by up to 30%^95^. Although SNAP is beneficial, one study found that even SNAP participants indicated that the cost of food was a major barrier to purchasing nutritious foods, and that SNAP payments did not support a healthy diet^61^. Findings from another study reported that food-insecure SNAP participants require approximately $10 to $20 more per person each week to buy sufficient food to meet their needs^86^. Therefore, expansion of benefits may promote food security among households with children.

Individuals from low-income households who participate in SNAP are also eligible to participate in SNAP-Education (SNAP-Ed), which is an evidence-based nutrition education program focusing on nutrition, budgeting, and a healthy lifestyle^87^. SNAP-Ed can play a significant role in improving dietary quality and food security^88^. In previous research, SNAP-Ed has been shown to improve participants’ food security^87,89,90,91^. However, SNAP-Ed, has not demonstrated improvements in participants’ dietary quality^88^. This review can help to shape future SNAP-Ed efforts to address nutritional quality. SNAP-Ed should continue to offer education to help participants optimize diet quality while maximizing the use of their food dollars. Along with these areas of focus, parental behavior and attitudes should be topics considered for future SNAP-Ed educational sessions in households with children. Given the relationship between parental behaviors and child dietary quality, educating parents on the importance of modeling healthy eating behaviors in the home and providing children with encouragement and support may support healthful eating. Promoting early exposure to a range of nutrient-dense foods, including fruits and vegetables, should be encouraged as it may improve diet quality in future life stages and promote positive attitudes towards eating throughout childhood and adolescence. Additionally, educating parents on the implications of negative feeding practices and parenting styles, such as pressure to eat and food restriction practices, on their child’s dietary quality, should also inform the development of educational lessons. Practical strategies to focus on include improving cooking skills, enhancing meal selection and planning knowledge, and improving attitudes toward healthy eating.

Most studies contained multiple themes illustrating their interrelationship. For example, if a parent purchases less nutritious food and brings it home, they increase the availability of these foods to their children and, as a result, build their food preference for these types of food. To obtain the greatest outcomes in terms of enhancing dietary quality and food security, components must be evaluated together, and all must be focused on during interventions.

Focusing on households with school children is important because the prevalence of food insecurity is disproportional among certain groups^96^. Additionally, the prevalence of food insecurity can vary within a household, in 7.8% of households with children, only the adults were affected by food insecurity. Parents often protect their child against food security by adapting the parents own intake^1^. Furthermore, race/ethnic status also impacts food security. In 2020, non-Hispanic black (21.7%) and Hispanic groups (17.2%) had a higher prevalence of food insecurity compared with the general population (10.5%)^97^. Therefore, it is important to recognize that factors may vary between households of different ethnicities, and these households may engage in behaviors that could vary from the ones featured in this review.

## 6 Limitations

Only one databsase was utilized in the search method for this review. Thus, relevant studies published in other databases may have been excluded. Furthermore, the search was refined to only include research published within the previous ten years, which may have resulted in the exclusion of high-quality, evidence-based studies.

While the goal of a scoping review is to be broad, exclusion criteria must be established. Research that did not focus on school-aged children were omitted. Therefore, studies that explored exclusively preschool-aged children or younger were removed. These studies may have offered additional relevant findings as some households with young children also include older children and may impact the overall dietary quality and food security in the household. Furthermore, environmental, community, and social variables that may have an influence on dietary quality and food security in families were not examined in this research.

Most studies included in this review were cross-sectional. This study design is not able to provide support for causal relationships. Longitudinal and potentially randomized experimental studies would be beneficial to provide stronger evidence.

## 7 Conclusion

Low income and food insecure households experience a variety of factors which impact the dietary quality of the food in the home. However, this review has identified several gaps within the literature that future studies could address. These gaps include further investigation of the associations between the factors, parental behaviors, child/adolescent behaviors, and dietary outcomes of children in these households over time. Longitudinal studies would be beneficial to strengthen the findings. The findings of this review can be used to inform the design and implementation of future nutritional education interventions in families with children aimed at improving dietary quality in low-income and food-insecure groups. From the above evidence, multiple factors influence the dietary quality and food security of low-income households with children in the U.S. and are reciprocally interlinked, and therefore must be considered collectively.

## Supporting information

PRISMA Scoping Review Checklist

## Data Availability

All data produced in the present work are contained in the manuscript.

## 8 Conflict of interest

The authors declare that the research was conducted in the absence of any commercial or financial relationships that could be construed as a potential conflict of interest.

## 9 Funding

This work was supported by US Department of Agriculture, National Institute of Food and Agriculture (USDA NIFA) (grant 2022-68015-36279), and USDA NIFA Hatch project (grant IND030489). The funders had no role in the design of the study; in the collection, analyses, or interpretation of data; in the writing of the manuscript, or in the decision to publish the results.

## Notes

### Competing Interest Statement

The authors have declared no competing interest.

